# Decidual-Like Natural Killer Cells Are Enriched in Melanomas and Acquire CD9 Through Melanoma Cell Contact

**DOI:** 10.64898/2026.09.18.26363442

**Authors:** Natalie A. Vandeven, Christopher E. Collora, Claire Muckle, Sofia Celli, Jessica S.W. Borges, Kylie Michel, Victoria Sorensen, Katherine Foley, Katherine M. Kichula, Martin D. McCarter, Sapna P. Patel, Kasey L. Couts, William A. Robinson, Richard P. Tobin, Paul J. Norman

## Abstract

Natural killer (NK) cells at the maternal-fetal interface harbor a specialized decidual phenotype that promotes placentation and immune tolerance. Although it has been postulated that tumors can exploit similar NK cell programs to augment the tumor microenvironment, this mechanism has not been investigated in melanoma. Using flow cytometry and multiplex immunohistochemistry, we identified CD56^Bright^CD16^−^CD49a^+^CD9^+^ decidual-like NK (dl-NK) cells within melanoma tumors; these cells were rare or absent in peripheral blood from either patients with melanoma or healthy donors. These dl-NK cells were detected within and surrounding melanoma tumors in both male and female patients and across melanoma subtypes and across primary, lymph node, and distant metastatic sites. In an exploratory cohort of patients treated with tumor-infiltrating lymphocyte therapy, higher dl-NK frequency was observed in the patient with progressive disease. Because CD9 is a defining feature of decidual NK cells, we examined the relationship of CD9 with the dl-NK phenotype. Tumor CD9 expression was positively associated with intratumoral dl-NK frequency (Pearson rho = 0.40, p = 0.056). Direct co-culture of healthy donor NK cells with CD9-expressing melanoma cells induced robust CD9 acquisition by CD56^Bright^ NK cells, whereas separation by a transwell abrogated this effect, demonstrating a requirement for direct cell-cell interaction. Collectively, these findings identify a dl-NK cell population within melanomas and suggest that direct interactions with melanoma cells may contribute to acquisition of this phenotype, revealing a potential immunologic parallel between the melanoma microenvironment and the maternal-fetal interface.

## INTRODUCTION

Melanoma is a lethal skin cancer with a rising incidence. In 2022 alone, nearly 100,000 new cases and >7,500 melanoma-related deaths were reported in the United States.^1,2^ Immune checkpoint inhibitors (ICIs) and more recently tumor-infiltrating lymphocyte (TIL) therapy have transformed treatment for patients with unresectable/metastatic disease.^3,4^ Additionally, ICI therapies are now being prescribed in the adjuvant and neoadjuvant settings for non-metastatic disease.^3,5,6^ Despite these advances, many patients experience severe ICI-related toxicities, and half either fail to respond in the first-line setting to ICI or in the second-line setting to TIL therapy.^3,4,7^ These limitations underscore a critical unmet need for biologically informed biomarkers that predict the risk of disease progression and immunotherapy response. Addressing this gap requires a mechanistic understanding of how melanomas *invade tissues* and *escape immune recognition*.

Although cancer has existed since the emergence of multicellular organisms, the capacity for malignant cells to invade tissues and evade immune surveillance may be a by-product of placentation (the formation of the placenta).^8–10^ Comparative studies reveal that mammals with non-invasive placentas develop melanoma at lower rates with limited metastatic capacity, whereas species with highly invasive placentas show increased melanoma incidence and metastatic potential.^10^ Humans possess the most invasive placentae, enabling extensive uterine spiral artery remodeling to support fetal growth and brain development.^11^ Strikingly, this same evolutionary adaptation is associated with disproportionately higher rates of metastatic melanoma, suggesting an evolutionary trade-off between reproductive success and cancer metastatic susceptibility.^10^

Successful placentation also depends on fetal cells evading maternal immune elimination within the maternal decidua (uterine lining), a process orchestrated by a specialized natural killer (NK) cell population called decidual NK (dNK) cells. These cells, defined by dual CD49a and CD9 expression, are functionally distinct from circulating NK cells: they exhibit limited cytotoxicity while actively promoting tissue invasion, angiogenesis, and immune tolerance.^12–14^ Decidual-like NK (dl-NK) cells which also express CD49a and CD9 have been described in several solid malignancies.^15–18^ In ovarian cancer, tumor-driven trogocytosis of CD9 has been implicated in facilitating dl-NK cell differentiation.^18^

In melanoma, tumor CD9 expression has been associated with worse disease-free survival^19^ and with adverse pathological features, including greater Breslow thickness, tumor ulceration, and sentinel lymph node positivity.^20^ Immunohistochemistry (IHC) staining of CD9 expression within melanoma tumors has shown localization around lymphatic and blood vessels, suggesting that CD9 may support melanoma invasion and metastasis.^20^ However, it remains unknown whether melanomas can also exploit tumor-driven trogocytosis of CD9 to promote dl-NK cell differentiation and foster a locally immunosuppressive tumor microenvironment similar to that of the maternal-fetal interface.

Here, we provide the first evidence, to our knowledge, that melanoma tumors harbor dl-NK cells both in male and female patients across primary and metastatic sites of disease and that melanoma cell lines can actively induce CD9 expression on NK cells through direct cell-cell contact. These findings suggest that melanomas may be co-opting an evolutionarily conserved pregnancy-associated immune program to facilitate tumor immune evasion. In addition, in a small exploratory cohort of patients treated with TIL therapy, higher intratumoral dl-NK frequency was observed pre-treatment in a patient with progressive disease. By identifying a previously unrecognized pregnancy-associated NK cell phenotype in melanoma, our work provides a framework for investigating how placentation-associated immune programs may contribute to melanoma immune escape

## RESULTS

### dl-NK cells are enriched within tumors but are rare in peripheral blood

To determine whether dl-NK cells are present in melanomas, we first compared NK cell populations in peripheral blood mononuclear cells from healthy donors (HD PBMC) and patients with melanoma (mel PBMC), and in melanoma tumor samples (mel tumor) using flow cytometry. NK cells were gated as shown in **Supplementary Figure 1** and separated into CD56^Dim^CD16^+^ (“CD56^Dim^”), CD56^Bright^CD16^+^ (“CD56^Int^”), and CD56^Bright^CD16^−^ (“CD56^Bright^”) populations (**Figure 1A–C**). Consistent with prior reports,^21,22^ CD56^Bright^ NK cells were the predominant intratumoral NK cell population (mean= 52.6% of total NK cell population; range 27.5% - 69.6%), whereas CD56^Dim^ NK cells predominated in HD PBMC (mean 79.3% total NK cells; range 66.6% - 89%) and mel PBMC (mean 81.9%; range 51.4% - 93.3%). A CD56^Int^ NK cell population was also enriched within mel tumors (mean 17.7% of total NK cells; range 8.01% - 27.9%) compared with mel PBMC (mean 1.76%; range 0.15% - 6.44%) and HD PBMC (mean 2.81%; range 0.062% - 5.64%) consistent with prior reports in melanoma.^23^

**Figure 1:**
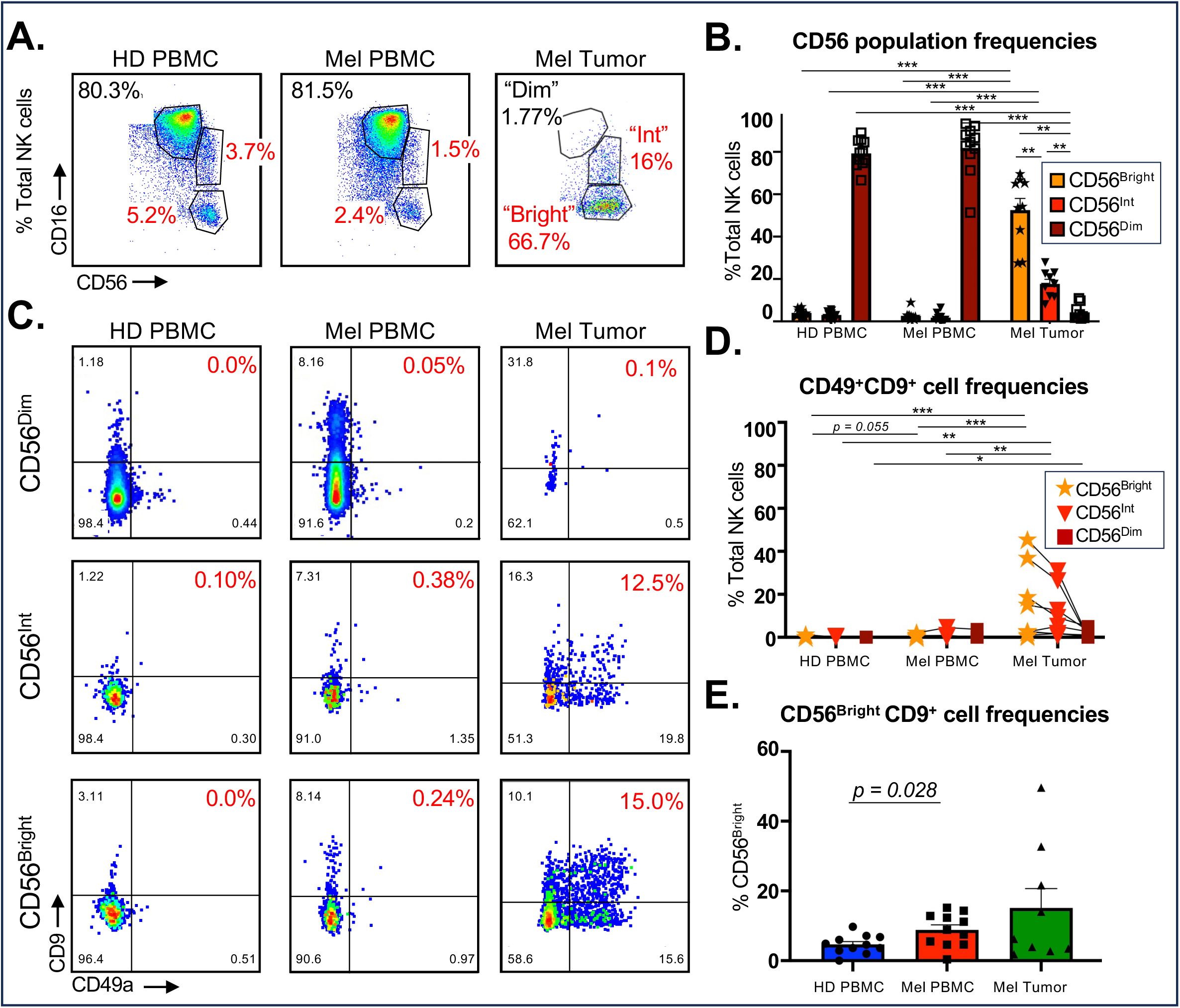
dl-NK cells are present within tumors but not peripheral blood. **(A)** shows representative plots of NK cells from healthy donors (HD; left), melanoma patient PBMC (Mel; middle) and melanoma patient tumors (right). CD56^Dim^CD16^+^ cells are denoted as “CD56^Dim^”; CD56^Bright^CD16+ cells as “CD56^Int^”; CD56^Bright^CD16^-^ cells as “CD56^Bright^”. **(B)** Summarized data from HD PBMC (n = 10), mel PBMC (n = 11), and mel tumors (n = 9). **(C)** shows representative plots of CD49a and CD9 staining across CD56^Dim^ (left), CD56^Int^ (middle) and CD56^Bright^ (right) NK cells. **(D)** shows summarized data from multiple donors with lines denoted matched samples**. (E)** shows summarized data of CD9 expression on CD56^Bright^ NK cells on HD PBMC (n = 10), mel PBMC (n = 11), and mel tumors (n = 9).

We next evaluated expression of CD49a and CD9, the canonical markers of dNK cells.^13^ We observed CD49a^+^CD9^+^ NK cells predominantly within the CD56^Bright^ population (consistent with a dl-NK phenotype), and these cells were markedly enriched in mel tumors (mean 13.9% of CD56^Bright^; range 0% - 45.4%) compared with mel PBMC (mean 0.47%; range 0% - 2.08%) or HD PBMC (mean 0%; range 0% - 1.21%; **Figure 1D**). When evaluating expression of CD9 in isolation, CD56^Bright^CD9^+^ cells were significantly enriched in mel PBMC (mean 8.87% of CD56^Bright^ NK cells; range 0.37% - 15.2%) compared with HD PBMC (mean 4.65%; range 0% - 9.73%; p = 0.028; **Figure 1E**). Together, these findings identify a distinct CD56^Bright^CD16^−^CD49a^+^CD9^+^ NK cell population within melanoma tumors and an enrichment of CD56^Bright^CD16^-^CD9^+^ NK cells within the peripheral blood of melanoma patients. However, it is important to note that here, tumor and peripheral blood samples were not matched from the same patients.

We next investigated whether the CD56^Int^ population more closely resembled CD56^Dim^ or CD56^Bright^ NK cells by assessing CXCR3, KIR2DL2/3/S2, and PD-1 expression (**Figure 2**). CXCR3, which is preferentially expressed on CD56^Bright^ NK cells and promotes NK cell infiltration into the tumor microenvironment,^24,25^ was expressed at higher levels on CD56^Bright^ than CD56^Dim^ NK cells across HD PBMC, mel PBMC, and mel tumors. The CD56^Int^ population showed a CXCR3 expression pattern more like CD56^Bright^ NK cells (**Figure 2A**).

**Figure 2.**
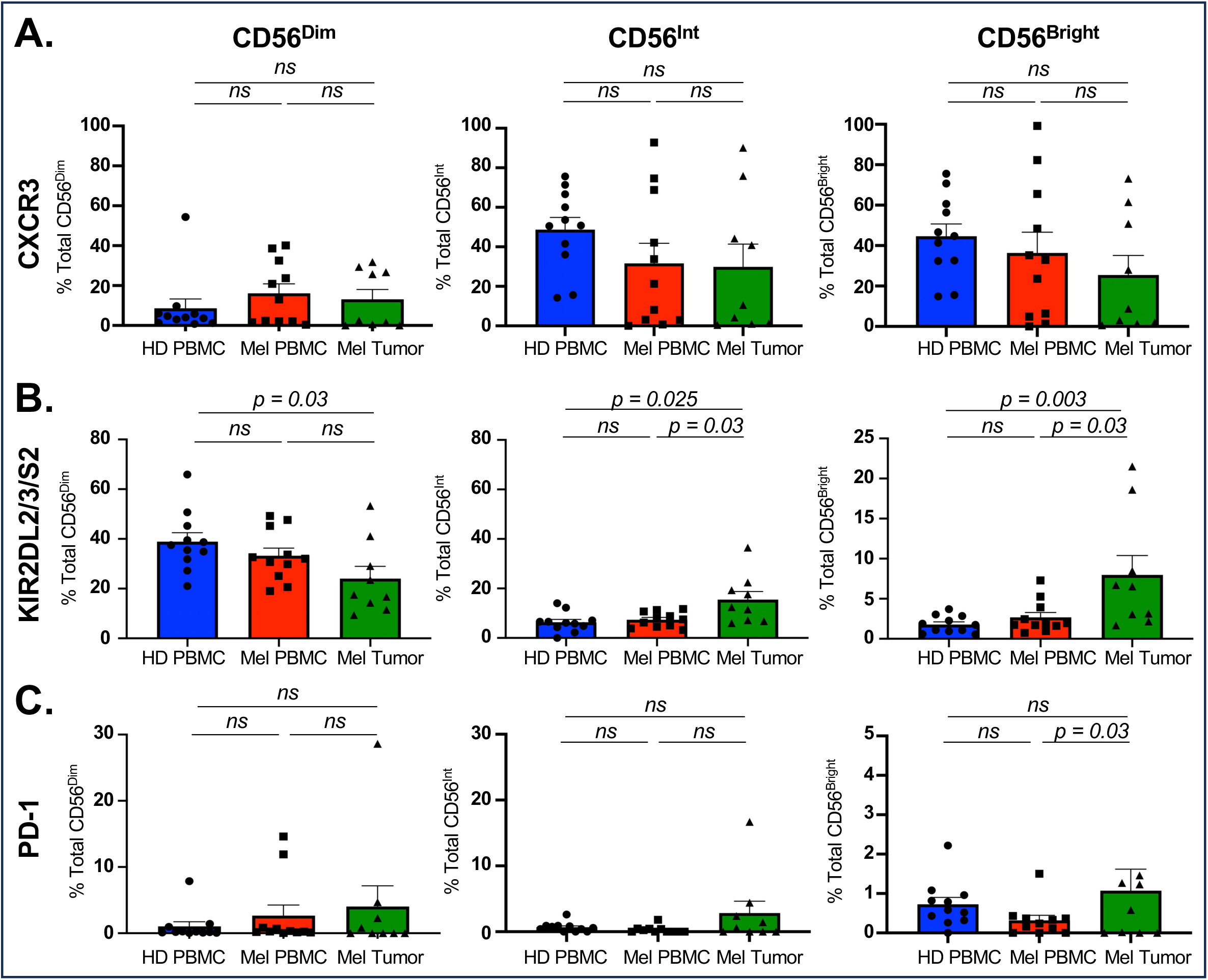
Expression of CXCR3, KIR2DL2/3/S2, and PD-1 across circulating and intratumoral NK-cell subsets in melanoma. Expression of **(A)** CXCR3, **(B)** KIR2DL2/3/S2, and **(C)** PD-1 was assessed within CD56^Dim^, CD56^Int^, and CD56^Bright^ NK cell populations from peripheral blood mononuclear cells (PBMCs) of healthy donors (HD PBMC), PBMCs of patients with melanoma (Mel PBMC), and melanoma tumor samples (Mel Tumor). Data are shown as the percentage of total NK cells within each indicated NK-cell subset expressing the respective marker. Each symbol represents an individual sample; bars represent mean ± SEM. Horizontal lines indicate pairwise statistical comparisons, with exact *P* values shown; *ns* denotes not significant.

KIR2DL2/3/S2 expression, which is predominantly associated with CD56^Dim^ NK cells,^26^ was higher on CD56^Dim^ than CD56^Bright^ NK cells across HD PBMC, mel PBMC, and mel tumors. However, KIR2DL2/3/S2 expression was significantly increased on intratumoral CD56^Int^ and CD56^Bright^ NK cells compared with their peripheral blood counterparts, whereas expression was decreased on intratumoral CD56^Dim^ NK cells (**Figure 2B**). Thus, the CD56^Int^ population again more closely resembled the CD56^Bright^ population.

Finally, PD-1 expression was low across NK cell subsets, consistent with its limited expression on NK cells,^27–29^ but was increased across intratumoral CD56^Dim^, CD56^Int^, and CD56^Bright^ populations compared with peripheral blood (**Figure 2C**). Collectively, these findings demonstrate remodeling of the intratumoral NK cell compartment and indicate that the CD56^Int^ subset more closely resembles CD56^Bright^ than CD56^Dim^ NK cells based on CXCR3, KIR2DL2/3/S2, and PD-1 expression.

### dl-NK cells are present within and adjacent to melanoma tumors

To confirm the presence and spatial distribution of dl-NK cells within intact melanoma tissue, we performed mIHC. Melanoma cells were identified by nuclear SOX10 expression, whereas dl-NK cells were characterized using CD56, CD16, CD49a, and CD9 (**Figure 3**). Cells exhibiting a CD56^+^CD16^−^CD49a^+^CD9^+^SOX10^−^ phenotype were identifiable within melanoma tumors (mean 5.24% of NK cells; range 0% - 20.6%) and peritumorally (mean 6.35%; range 1.2% - 16.7%), confirming the presence of dl-NK cells using an orthogonal tissue-based approach. Representative images demonstrated dl-NK cells within the tumor microenvironment and in regions surrounding tumor cells (**Figure 3**). These populations were detected across primary tumors, lymph nodes, and distant metastatic sites and in tumors from both female and male patients (**Supplementary Figure 2**).

**Figure 3:**
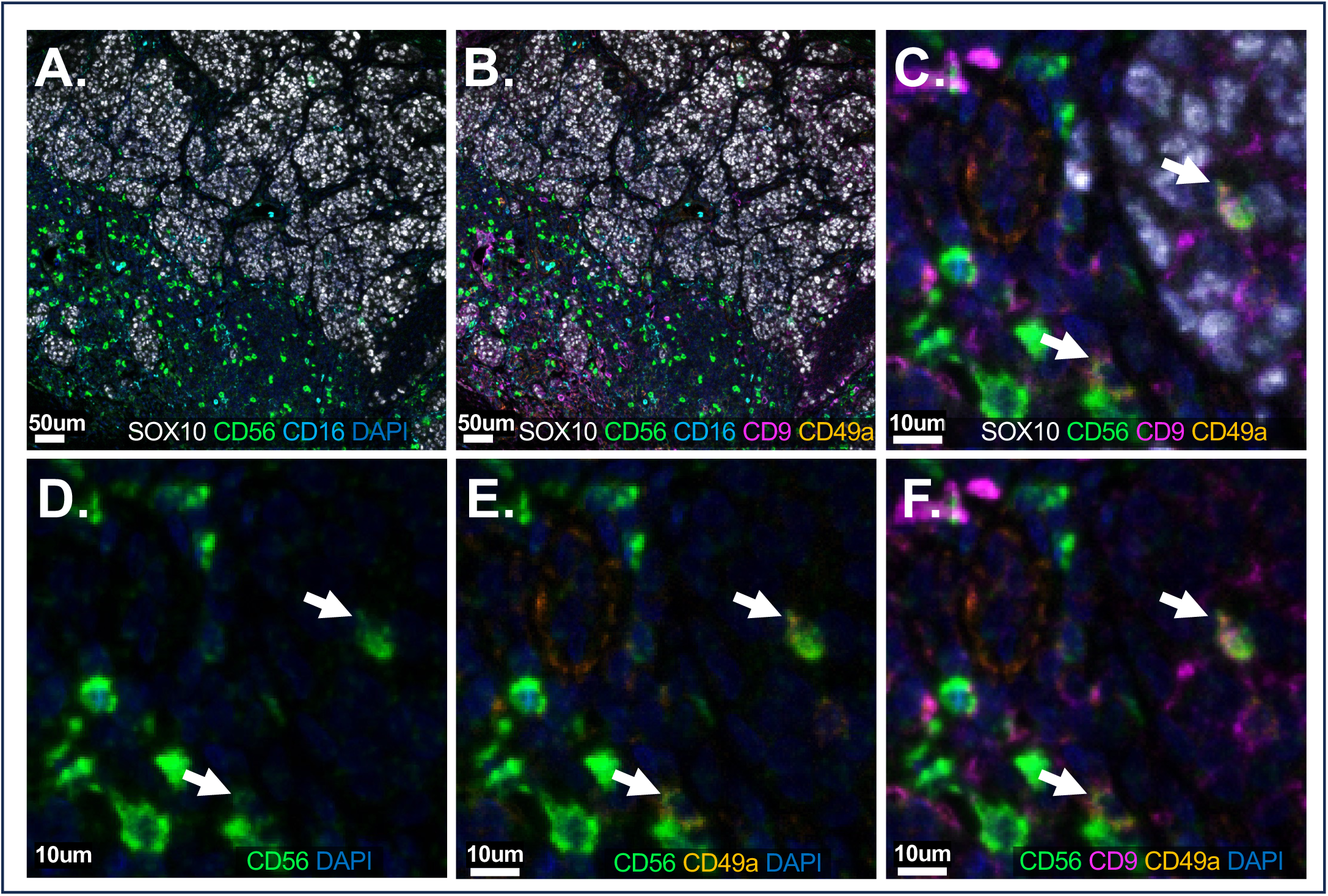
dl-NK cells are present within tumors and peritumorally in men and women. A-F: show representative images of multiplexed immunohistochemistry slides. **(A)** shows tumor markers staining positive for SOX10 (white), NK cells staining with CD56 (green) and CD16 (cyan). **(B)** shows the same tumor section with the addition CD9 (magenta) and CD49a (yellow) staining to identify dl-NK cells (defined as CD56^+^CD16^-^CD9^+^CD49a^+^SOX10^-^). **(C)** shows two dl-NK cells (white arrows) at higher magnification. **(D)** depicts the two dl-NK cells with CD56 (green) and DAPI channels only. **(E)** shows the two dl-NK cells with CD56, CD49a (yellow) and DAPI. **(F)** Shows the two dl-NK cells with CD56, CD49a, CD9 (pink) and DAPI

### Intratumoral dl-NK cells are detected across melanoma subtypes and clinical characteristics

We next investigated whether intratumoral dl-NK frequency differed according to melanoma subtype or patient and tumor characteristics across the entire cohort, including those assessed using flow cytometry and mIHC (**Figure 4**). Although this exploratory cohort predominantly consisted of patients with cutaneous melanoma, dl-NK cells were detected across cutaneous (n = 25), mucosal (n = 3), acral (n = 3), uveal (n = 1), and unknown-primary melanomas (n = 1), with no significant difference in frequency among melanoma subtypes (Kruskal-Wallis *P*=0.88; **Figure 4A**).

**Figure 4.**
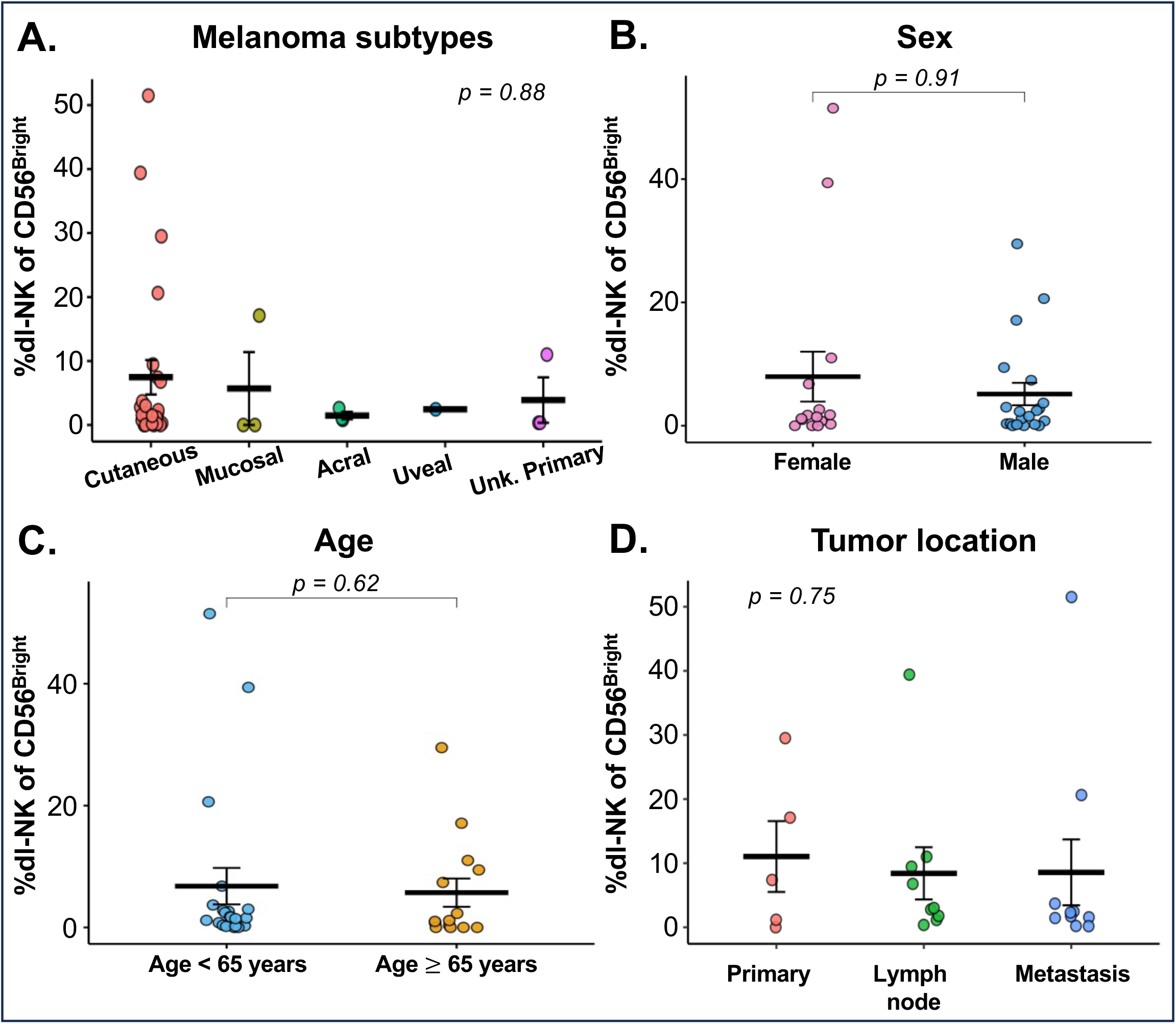
dl-NK cells detected across melanoma subtypes and patient and tumor characteristics. Intratumoral dl-NK cell frequency was compared across melanoma subtypes and clinical characteristics. **(A)** dl-NK frequency according to melanoma subtype, including cutaneous, mucosal, acral, uveal, and unknown-primary melanoma. Differences among melanoma subtypes were assessed using the Kruskal–Wallis test. **(B)** dl-NK frequency according to sex. **(C)** dl-NK frequency according to age at sample collection, stratified by age < 65 years versus ≥ 65 years. Differences between two groups in panels B and C were assessed using the Wilcoxon rank-sum test. **(D)** dl-NK frequency according to tumor sampling site, categorized as primary tumor, lymph node, or other metastatic site. Differences among tumor sites were assessed using the Kruskal–Wallis test. Individual points represent individual patient samples; horizontal bars and error bars indicate mean ± SEM. *P* values are shown for each comparison.

Similarly, dl-NK frequency did not significantly differ according to sex (female, n = 15; male, n = 20; Wilcoxon p = 0.91; **Figure 4B**) or age at sample collection (<65 years, n = 21; ≥65 years, n = 14; *p* = 0.62; **Figure 4C**). dl-NK cells were also detected in primary tumors (n = 5), lymph nodes (n = 9), and other metastatic sites (n = 10), with no significant difference in frequency according to tumor sampling site (Kruskal-Wallis *p =* 0.75; **Figure 4D**). Thus, dl-NK cells were detected across melanoma subtypes, sexes, age groups, and primary, lymph node, and other metastatic sites, with no significant differences in frequency among the groups examined.

### dl-NK frequency and tumor CD9 expression vary among patients undergoing TIL therapy

To explore the relationship between dl-NK cells and treatment response, we analyzed matched tumor and peripheral blood samples from four patients with cutaneous melanoma who had progressed on three prior lines of therapy and subsequently received TIL therapy (**Figures 5 and 6**). Tumor tissue was obtained from tissue remaining after tumor procurement for TIL manufacturing, and PBMC was collected from the same patients on the date of tumor tissue collection (pre-treatment).

**Figure 5.**
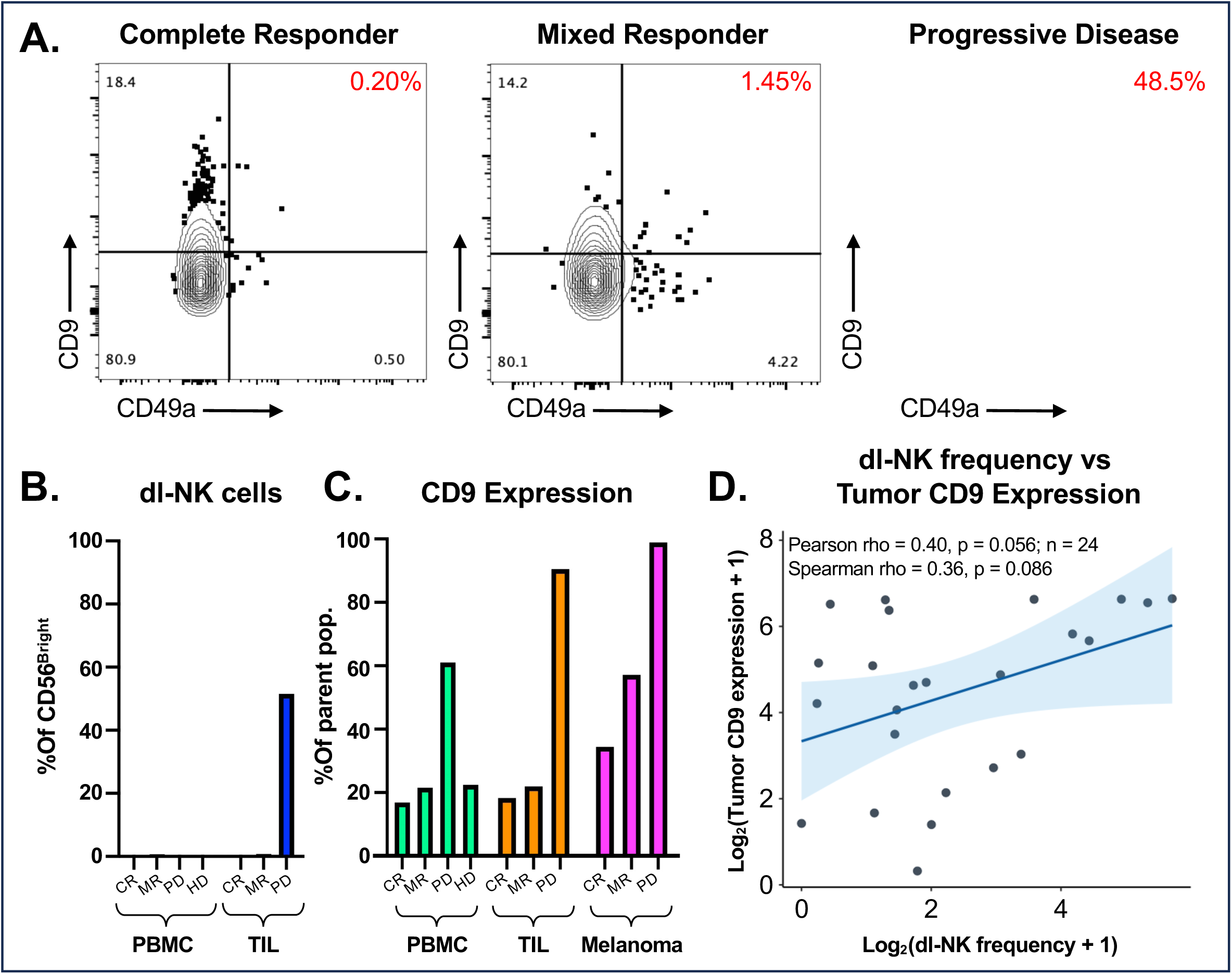
Decidual-like NK (dl-NK) cell phenotype and tumor CD9 expression according to clinical response to treatment with tumor-infiltrating lymphocyte (TIL) therapy. **(A)** Representative flow cytometry plots showing CD49a and CD9 expression on intratumoral NK cells from patients who had previously failed ICI therapy and were subsequently received TIL therapy and achieved a complete response (CR), mixed response (MR), or progressive disease (PD) per RECIST criteria. The frequency of CD49a⁺CD9⁺ dl-NK cells is indicated in red. **(B)** dl-NK cell frequency in peripheral blood (PBMC) and intratumoral (TIL) CD56^Bright^ NK cell populations stratified by clinical response. Within the peripheral blood CD9 expression on patient samples was also compared to CD9 expression on CD56^Bright^ NK cells from healthy donors (HD). **(C)** CD9 expression across peripheral blood NK cells (PBMC), intratumoral NK cells (TIL) and on melanoma tumor cells (melanoma) stratified by clinical response. CD9 expression is shown as the percentage of the indicated parent population. **(D)** Association between intratumoral dl-NK frequency and tumor CD9 expression. Both variables were log₂-transformed as log₂(value + 1). The solid line represents the linear regression fit, with the shaded region indicating the 95% confidence interval. Tumor CD9 expression showed a positive trend with dl-NK frequency (Pearson *rho* = 0.40, *p* = 0.056; Spearman ρ = 0.36, *p* = 0.086; *n* = 24).

**Figure 6.**
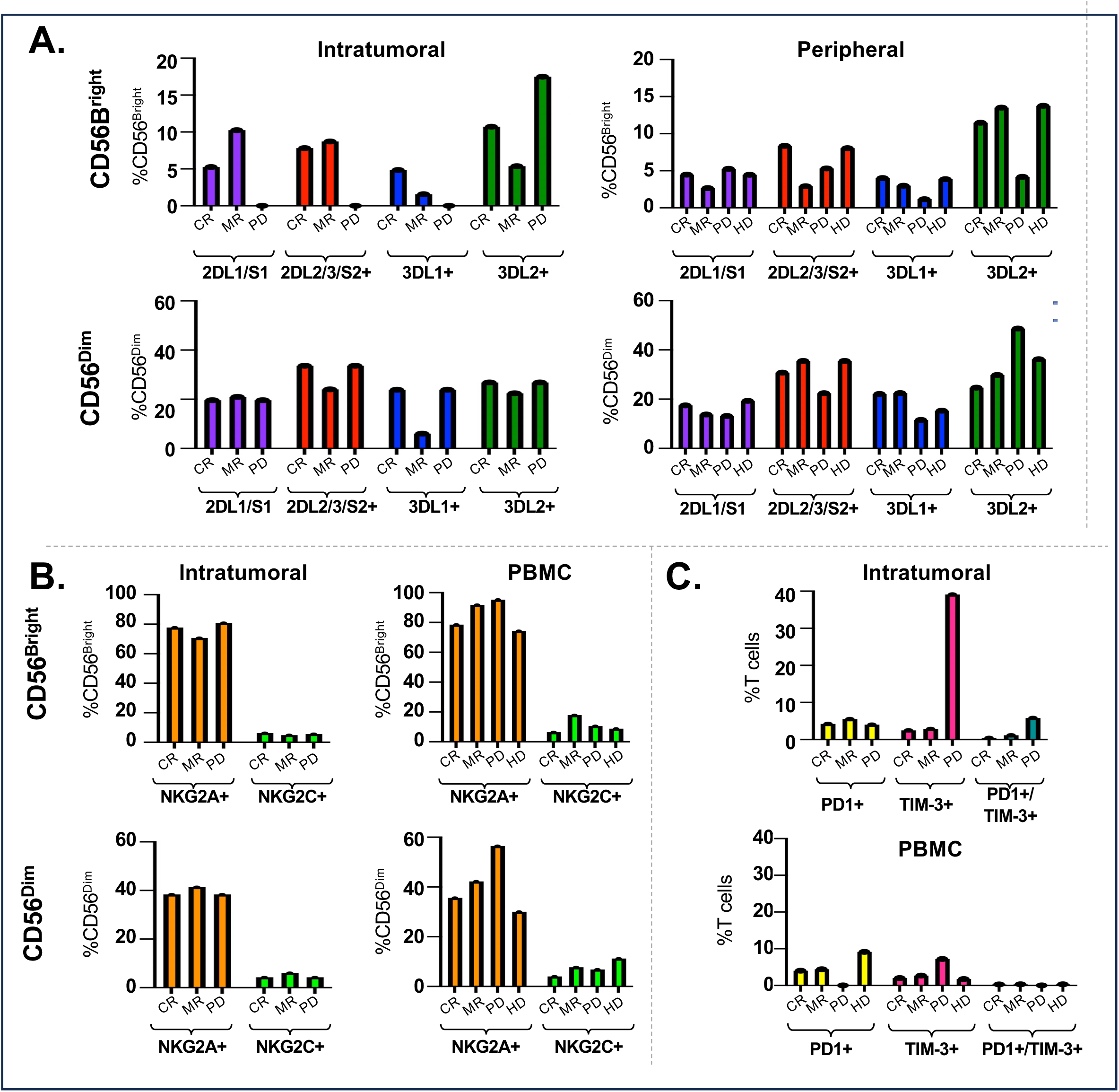
Phenotypic characterization of intratumoral and peripheral NK and T cells according to clinical response to TIL therapy. Immune-cell phenotypes were assessed in tumor-infiltrating lymphocytes (TIL) and matched peripheral blood mononuclear cells (PBMCs) and compared across patients with complete response (CR; n = 1), mixed response (MR; n = 2), progressive disease (PD; n = 1) to therapy or healthy donor (HD; n = 5). **(A)** Expression of killer cell immunoglobulin-like receptors (KIRs) on CD56^Bright^ and CD56^Dim^ NK cell populations in intratumoral and peripheral compartments. NK cells were grouped according to expression of KIR2DL1/S1, 2DL2/3/S2, 3DL1, and 3DL2. **(B)** Expression of NKG2A and NKG2C on CD56^Bright^ and CD56^Dim^ NK cells in intratumoral and peripheral compartments. **(C)** Frequencies of PD-1⁺, TIM-3⁺, and double positive PD-1⁺TIM-3⁺ T cells within intratumoral and peripheral T-cell populations. Bar height represents the percentage of the indicated parent population.

Flow cytometric analysis demonstrated variation in the frequency of dl-NK cells within metastatic tumor samples (i.e. the starting cell product for generating TIL). A tumor that progressed showed the highest dl-NK cell frequency (%dl-NK of CD56^Bright^ = 48.5%; PD; **Figure 5A & B**). Patients with tumors showing mixed response had considerably lower dl-NK cell frequencies (%dl-NK of CD56^Bright^ = 0.18% - 1.45%; MR; n = 2 plot shows results averaged), similar to a patient whose tumor completely responded (%dl-NK of CD56^Bright^ = 0.2%; CR; **Figure 5A & B**). Within the peripheral blood, dl-NK cells were largely undetectable across all patients and age-and sex-matched healthy donor controls (n = 5; **Figure 5B**).

We next evaluated CD9 expression across peripheral and intratumoral NK cell populations and on melanoma tumor cells (**Figure 5C**). The patient with PD did not have detectable circulating dl-NK cells (%dl-NK of CD56^Bright^ = 0.29%) but had high CD9 expression on CD56^Bright^ NK cells both within the peripheral (%CD9^+^ of CD56^Bright^ = 61%) and intratumoral compartments (%CD9^+^ of CD56^Bright^ = 90.7%), as compared with patients with CR (peripheral %CD9^+^ of CD56^Bright^ = 16.9% and tumoral 18.4%) or MR (peripheral %CD9^+^ of CD56^Bright^ = 21.5% and tumoral %CD9^+^ of CD56^Bright^ = 22.0%) and HD (%CD9^+^% of CD56^Bright^ = 22.5%; **Figure 5C**). In addition, melanoma tumor CD9 expression was high in the patients with MR (%tumor cells CD9^+^ = 57.3%) and PD (%tumor cell CD9^+^ = 99.1%), whereas the complete responder had comparatively lower tumor CD9 expression (%tumor cells CD9^+^ = 34.5%; **Figure 5C**). Given the limited cohort size, these observations were considered exploratory.

Because CD9 is a defining feature of the dl-NK phenotype^13^, we next examined the relationship between tumor CD9 expression and intratumoral dl-NK frequency across the entire set of samples assessed by flow cytometry and mIHC (**Figure 5D**). Tumor CD9 expression showed a positive association with dl-NK frequency following log2 transformation (Pearson *r* = 0.40, p = 0.056; n = 24). A directionally similar relationship was observed using Spearman rank correlation (ρ = 0.36, p = 0.086; **Figure 5D**).

### Tumor and blood NK-and T-cell phenotypes vary by TIL therapy response

We further characterized NK and T cell phenotypes in matched pre-treatment tumor and PBMC samples from patients with CR, MR or PD following TIL therapy and compared peripheral profiles with healthy donor controls (**Figure 6**).

Expression of KIR2DL1/S1, KIR2DL2/3/S2, KIR3DL1, and KIR3DL2 varied across CD56^Bright^ and CD56^Dim^ NK cell populations and between intratumoral and peripheral compartments (**Figure 6A**). The greatest differences were observed within the intratumoral CD56^Bright^ compartment, which generally demonstrated decreased expression of KIR2DL1/S1, KIR2DL2/3/S2, and KIR3DL1 and increased expression of KIR3DL2 (**Figure 6A**). KIR genotypes were not available for these patients.

CD94/NKG2A and NKG2C expression did not demonstrate major differences within the intratumoral compartment (**Figure 6B**). However, the peripheral CD56^Dim^ NK cell population from the patient with PD exhibited increased NKG2A expression (%NKG2A^+^ of CD56^Dim^ = 56.5%) relative to the MR patients (average NKG2A^+^% of CD56^Dim^ = 42.2%), the CR patient (%NKG2A^+^ of CD56^Dim^ = 35.6%) and healthy donors (average NKG2A^+^% of CD56^Dim^ = 30.0%; **Figure 6B**).

We also examined the inhibitory receptors PD-1 and TIM-3 on T cells (**Figure 6C**). A higher frequency of TIM-3 mono-expression was observed particularly within the intratumoral compartment in the patient with PD (%TIM-3^+^ of total T cells = 39.1%) as compared to the patient with MR (tumoral %TIM-3^+^ of total T cells = 2.86%) and CR (%TIM-3^+^ of total T cells = 2.47%). A modest increase in dual PD-1 and TIM-3 expression was also observed within the intratumoral compartment of the patient with PD (%PD-1+TIM-3+ of total T cells = 5.8%), as compared to the patients with MR (average %PD-1^+^TIM-3^+^ of total T cells = 1.14%) or the patient with CR (%PD-1^+^TIM-3^+^ of total T cells = 0.46%) although overall these cells represented <10% of total T cells. Collectively, NK and T cell phenotypes varied among patients with different responses to TIL therapy.

### Direct interaction with CD9-expressing melanoma cells induces CD9 acquisition by CD56^Bright^ NK cells

Given the observed relationship between melanoma tumor CD9 expression and intratumoral dl-NK frequency, we next investigated whether direct interaction with melanoma cells could induce CD9 acquisition by NK cells as has been previously reported in ovarian cancer.^18^ CD56^Bright^ NK cells from healthy donors were cultured alone, stimulated with PMA/ionomycin, or co-cultured with melanoma cell lines expressing varying levels of CD9. Co-cultures were performed either in direct contact or using a transwell system to distinguish contact-dependent from soluble-factor-mediated effects (**Figure 7**).

**Figure 7:**
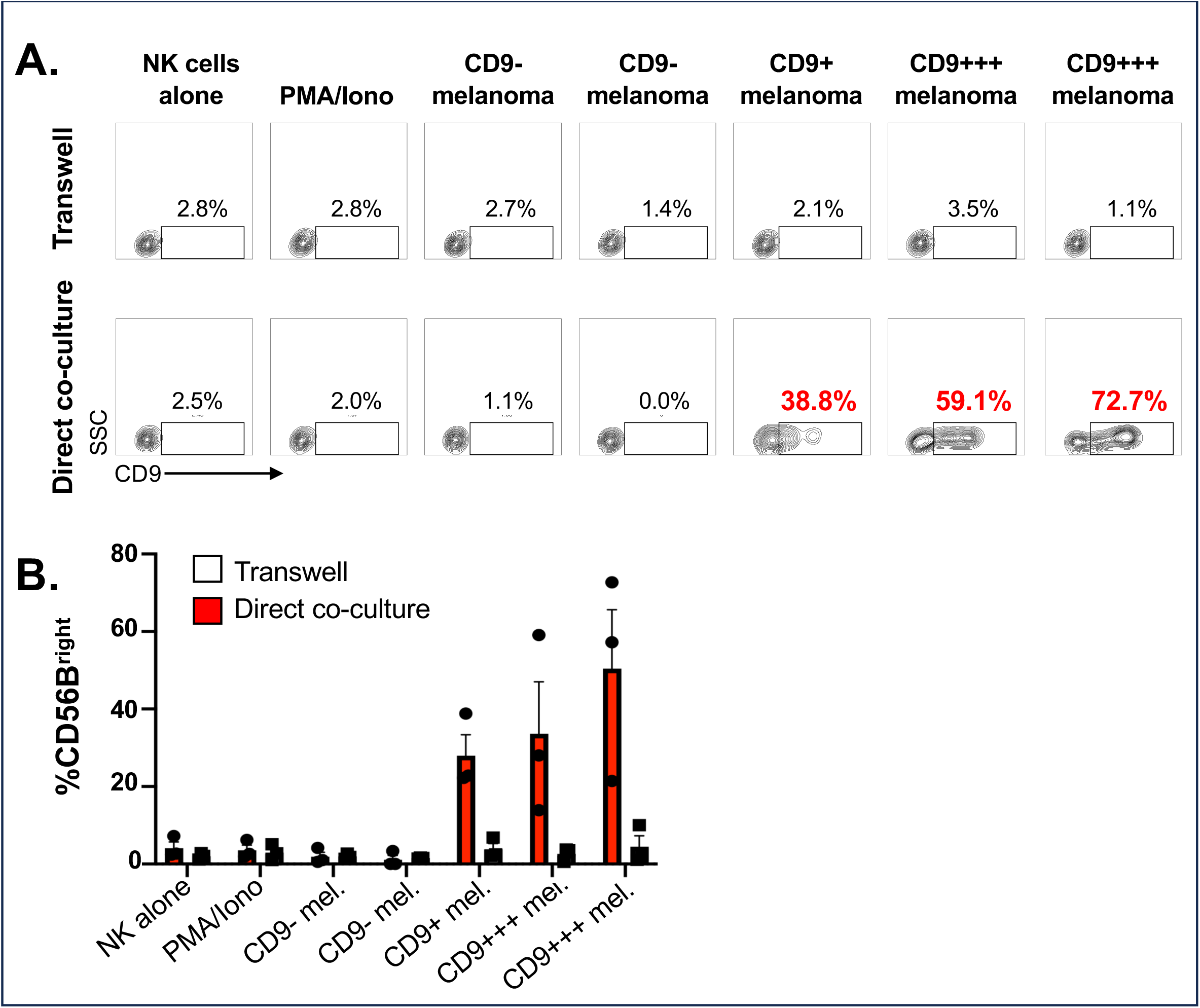
CD56^Bright^ NK cells acquire CD9 through direct co-culture with CD9^+^ melanoma cell lines. **(A)** shows representative flow plots from NK cells cultured under the listed conditions either in a transwell system (top row) or in direct co-culture (bottom row). Gated regions indicate the percentage of CD9^+^ cells within the CD56^Bright^ NK cell population. (**B)** Summary of these findings across three healthy donors; error bars represent mean ± SEM.

Direct co-culture with melanoma cells expressing a range of CD9 (details of cell line CD9 expression shown in **Supplementary Figure 3**) resulted in a significant increase in CD9 expression on CD56^Bright^ NK cells (ranging from 38.8% - 72.7%; **Figure 7A**), whereas NK cells cultured in a transwell system did not demonstrate comparable CD9 acquisition. CD9 expression remained low when NK cells were cultured alone or with CD9-negative melanoma cells (**Figure 7A**). These data remained consistent when testing CD9 acquisition across three healthy donors (**Figure 7B**). Thus, robust CD9 acquisition by CD56^Bright^ NK cells required direct contact with CD9-expressing melanoma cells under these experimental conditions.

## Discussion

In this study, we identified a population of CD56^Bright^CD16^-^CD49a^+^CD9^+^ dl-NK cells that were enriched within melanoma tumors and by contrast were rare in the peripheral blood of patients with melanoma or healthy donors. These cells were identified using both flow cytometry and mIHC and were observed across melanoma subtypes and in both male and female patients. Furthermore, direct co-culture with CD9-expressing melanoma cells resulted in robust acquisition of CD9 by CD56^Bright^ NK cells, whereas separation of NK cells from melanoma cell lines using a transwell system largely abrogated this effect. Collectively, these findings support a model in which melanoma cells can locally reshape NK cell phenotype through direct cell-cell interactions and raise the possibility that melanoma co-opts elements of an immune program normally operating at the maternal-fetal interface.

NK cells at the maternal-fetal interface are phenotypically and functionally distinct from conventional circulating NK cells. Rather than functioning primarily as cytotoxic effectors, dNK cells participate in trophoblast invasion, vascular remodeling, and maintenance of local immune homeostasis during placentation.^17^ CD56^Bright^CD16^-^dNK cells express tissue-associated markers including CD49a and CD9 and interact extensively with invading extravillous trophoblasts and other decidual cells.^17^ CD49a is an integrin that regulates migration, retention and preservation of immune cells.^30^ CD9 likely regulates integrin cell-surface organization, proliferation, adhesion and signaling;^31^ together, their expression is associated with tissue residency and the pro-angiogenic phenotype of dNK cells. This specialized phenotype provides a compelling biological analogy to cancer: both placentation and malignant progression require tissue invasion, vascular remodeling, and local modulation of immune surveillance. Prior comparative and evolutionary studies have proposed that mechanisms enabling invasive placentation and mechanisms underlying malignant invasion are biologically interconnected: an evolutionary framework that motivated the present study. The identification of dl-NK cells in melanoma extends observations from other solid tumors demonstrating that tumor-associated NK cells can acquire features normally associated with dNK cells. CD56^Bright^CD16^-/low^CD49a^+^CD9^+^ NK populations have been described in lung, colorectal, and ovarian cancers and have generally been associated with reduced cytotoxic and/or proangiogenic functions.^16^ In colorectal cancer, for example, tumor-associated and tumor-infiltrating NK cells exhibit a dl-NK cell phenotype accompanied by impaired degranulation and production of factors capable of promoting endothelial proliferation, migration, and capillary-like structures.^16^ Thus, tumor-associated NK cells may not simply represent ineffective antitumor lymphocytes but, under some conditions, may be actively reprogrammed toward functions that support the tumor microenvironment. Our identification of CD49a^+^CD9^+^ NK cells within melanoma tumors adds melanoma to this emerging paradigm.

The predominance of the dl-NK phenotype within the CD56^Bright^ compartment is particularly relevant given previous observations regarding NK cell biology in melanoma. Whereas CD56^Dim^ NK cells represent the dominant circulating population and generally possess greater direct cytotoxic capacity, CD56^Bright^ NK cells have stronger cytokine-producing and immunoregulatory properties. In melanoma, increased circulating CD56^Bright^ NK cell frequency has previously been associated with shorter progression-free and overall survival.^32^ CD56^Bright^ NK cells from patients with advanced melanoma also demonstrated altered functional responses, including reduced TNFα and GM-CSF production following stimulation.^32^ Our findings provide a potential additional layer of complexity to these observations by demonstrating that the CD56^Bright^ compartment within melanoma tumors contains cells expressing the canonical decidual-associated markers CD49a and CD9. Consistent with these observations, we found increased CD9 expression among circulating CD56^Bright^ NK cells in patients with melanoma compared with healthy donors, although fully defined CD49a^+^CD9^+^ dl-NK cells remained rare in peripheral blood. Whether the adverse clinical associations previously attributed to CD56^Bright^ NK cells are specifically related to expansion or induction of this dl-NK population remains unknown and warrants investigation in larger clinically annotated cohorts.

Our broader phenotypic characterization also suggests remodeling of NK cell receptor expression in melanoma. The CD56^Int^ population generally resembled CD56^Bright^ cells in CXCR3, KIR2DL2/3/S2, and PD-1 expression, suggesting that a transitional population between CD56^Dim^ and CD56^Bright^ NK cells may exist, although this cannot be established from these data alone. Differences in KIR2DL2/3/S2 expression were also observed between circulating and intratumoral NK populations; however, the combined KIR2DL2/3/S2 measurement does not distinguish inhibitory KIR2DL2/3 from activating KIR2DS2, limiting interpretation of these findings.

One of the most intriguing findings of this study is the relationship between tumor CD9 expression, CD9 expression on peripheral NK cells and the dl-NK phenotype. Across patient samples, tumor CD9 expression showed a positive association with intratumoral dl-NK frequency, although this relationship did not reach conventional statistical significance in this relatively small cohort (Pearson *r*=0.40, *p* = 0.056). CD9 itself has potentially important biological implications in melanoma. Although CD9 has been reported to suppress metastasis development in some malignancies^33^, its role is highly context dependent. In primary cutaneous melanoma, CD9 re-expression has been reported in more locally advanced melanomas, particularly within tumor-cell clusters located near or within lymphatic and blood vessels. CD9 tumor positivity was associated with sentinel lymph-node involvement, distant metastasis, and shorter melanoma-specific survival.^20^ These observations raise the possibility that CD9 expression by melanoma cells could have dual consequences: contributing directly to tumor invasion or dissemination while simultaneously influencing neighboring immune cells.

Our co-culture experiments provide initial mechanistic support for the latter possibility. CD56^Bright^ NK cells acquired high levels of CD9 following direct contact with CD9-expressing melanoma cells, while CD9 acquisition was largely absent when the same cells were physically separated by a transwell. By contrast, direct co-culture with CD9-negative melanoma cells did not induce the same phenotype. These findings closely parallel observations in high-grade serous ovarian carcinoma, in which NK cells can acquire tumor-derived CD9 through trogocytosis.^18^ Importantly, tumor-derived CD9 was not simply a phenotypic marker in that system: CD9 acquisition reduced NK cell antitumor cytokine production and cytotoxicity, whereas CD9 blockade or tumor CD9 knockout restored cytotoxic function.^18^ Together with our patient data, these findings raise the possibility that melanoma cells promote contact-dependent acquisition of CD9 by neighboring NK cells, potentially through a mechanism such as trogocytosis described in ovarian cancer.^18^ Together, these findings raise the possibility that NK cells acquire CD9 through contact-dependent mechanisms such as trogocytosis, generating a transitional circulating CD9^+^ NK cell population that subsequently localizes to tumors and acquires CD49a to adopt a dl-NK phenotype.^34^ If validated, circulating CD9^+^ NK cells could serve as a potential biomarker of intratumoral dl-NK cells

The spatial distribution of dl-NK cells may also provide insight into their biology. Multiplex immunohistochemistry identified these cells in both tumoral and peritumoral regions, and they were detected in primary tumors, involved lymph nodes, and distant metastases. Their presence across multiple sites is consistent with either recruitment of a susceptible NK cell population followed by local reprogramming or differentiation of tissue-resident NK cells in response to the tumor microenvironment. The contact dependence observed in our co-culture system favors a model in which at least part of the phenotype is acquired locally after NK cells encounter melanoma cells. Nevertheless, soluble factors known to influence NK cell polarization-including TGFβ, hypoxia-associated signals, and other immunoregulatory mediators-may cooperate with cell-contact-dependent mechanisms to establish and maintain the complete dl-NK state.^35^

The potential clinical significance of dl-NK cells is suggested by our exploratory analysis of patients undergoing TIL therapy. Among four heavily pretreated patients for whom matched tumor and peripheral blood were available, the tumor that failed to respond to TIL therapy demonstrated the greatest intratumoral dl-NK frequency together with high CD9 expression on tumor cells and CD56^Bright^ NK cells, whereas the complete responder had relatively low dl-NK and tumor CD9 expression. We also noted differences in NKG2A expression on peripheral CD56^Dim^ NK cells, with the nonresponding patient having notably higher NKG2A and lower KIR2DL2/3/S2 expression. Circulating CD56^Bright^ NK cells are typically NKG2A^+^ with low KIR expression. In contrast, progressive differentiation within the circulating CD56^Dim^ compartment is associated with decreasing NKG2A and increasing KIR expression.^36,37^ The pattern observed in the nonresponding patient could reflect differences in NK cell differentiation state or subset composition, but these cross-sectional data cannot establish receptor loss or reacquisition or functional inhibition. Given the small sample size, these observations cannot establish an association between dl-NK cells and treatment response and should not be interpreted as evidence that dl-NK cells mediate resistance to TIL therapy. Rather, they provide a hypothesis for prospective testing. Larger cohorts with pretreatment tissue, standardized clinical endpoints, and longitudinal sampling are needed to determine whether dl-NK frequency or tumor CD9 expression predicts response to TIL therapy, immune checkpoint blockade, or other immunotherapies.

Several limitations of this study should be acknowledged. First, the clinical cohorts are small and heterogeneous with respect to melanoma subtype, disease site, and treatment history, thereby limiting conclusions regarding clinical outcomes and subgroup differences. The absence of differences in dl-NK frequency according to sex, age, melanoma subtype, or tumor location should therefore not be interpreted as evidence of equivalence between these groups. Second, although CD56^Bright^CD16^-^CD49a^+^CD9^+^ cells phenotypically resemble decidual NK cells, the present study does not establish that they are functionally equivalent to uterine dNK cells. Transcriptional, epigenetic, and functional characterization will be required to determine the extent to which melanoma-associated dl-NK cells recapitulate the functional program of bona fide dNK cells. Third, our co-culture experiments establish a requirement for direct melanoma-NK interaction for robust CD9 acquisition but do not yet definitively identify trogocytosis as the mechanism. Fourth, the functional consequences of CD9 acquisition on primary human NK cells and the role of dl-NK cells within melanomas remain unknown. Finally, the absence of paired KIR genotyping and HLA ligand information limits interpretation of receptor-expression patterns.

These limitations also define several important directions for future investigation. Determining whether CD9 acquisition is necessary or sufficient for functional NK cell reprogramming will be highly informative. Experiments using CD9-deficient melanoma cells, CD9-blocking antibodies, and direct tracking of membrane transfer could establish the mechanism of CD9 acquisition, while paired cytotoxicity, cytokine, and angiogenesis assays could determine its functional consequences. Single-cell transcriptomic and spatial approaches could further determine whether dl-NK cells represent a discrete NK cell state, a transitional phenotype, or part of a broader continuum of tumor-induced NK cell differentiation and/or dysfunction. Finally, integrating NK cell phenotype with KIR genotype, HLA ligand expression, and clinical outcomes could identify the receptor–ligand interactions that permit melanoma cells to establish this phenotype and determine whether this axis represents a therapeutically actionable mechanism of immune escape.

In conclusion, our findings identify a CD56^Bright^CD16^-^CD49a^+^CD9^+^ dl-NK cell population within melanoma and provide evidence that melanoma cells can induce a defining component of this phenotype through direct cell-cell interaction. The striking parallel with the maternal-fetal interface raises the possibility that melanoma exploits an evolutionarily conserved program normally used to permit controlled tissue invasion and immune tolerance during placentation. Whether these phenotypic changes impair antitumor immunity remains to be determined. Defining the mechanisms and functional consequences of this reprogramming may uncover biomarkers of immune resistance and new strategies for restoring NK cell antitumor activity.

## METHODS

### Human Tissues

Healthy donor PBMC were obtained for the Human Immune Tissue Network Biobank by the University of Colorado Clinical and Translation Research Center (CTRC), in sodium heparin tubes under the University of Colorado COMIRB #17-2159. PBMC were isolated using a Ficoll gradient (Cytiva). In addition, plateletpheresis leukoreduction filters (LRS chambers) were purchased from Vitalant Blood Center (Denver, CO, USA). Melanoma blood and tumor samples were collected from the University of Colorado Hospital under the Melanoma Biorepository at the University of Colorado Cancer Center COMIRB #05-0309. Tumors were first mechanically dissociated into pieces < 1mm^3^ and then chemically digested at 37°C for 30 minutes with 5 µg mL−1 Liberase DL (Roche). Following enzymatic digestion, tumors were passed through a 40 μm cell strainer to create a single-cell suspension. Melanoma cell lines were generously donated by the Melanoma Biorepository and maintained in RPMI 1640 containing HEPES supplemented with 10% FBS, 100 U/mL penicillin, 100 μg/mL streptomycin at 37°C in a humidified 5% CO_2_ atmosphere.

### Flow cytometric analysis

Cells from tumors and PBMCs were stained in flow staining buffer containing 5% BSA and 0.05 M EDTA for all extracellular staining. A full list of flow cytometric antibodies can be found in **Supplementary Table 1**. All cells were stained with Live/Dead discrimination Fixable Viability Dye 780 (65-0865-14, eBiosciences). Cells were fixed using 1X BD Cytofix/Cytoperm solution (554723, BD Biosciences) for 20 min at 4°C. Flow cytometric analysis was performed using a Cytek Aurora spectral flow cytometer, and data were analyzed using FlowJo v10 Software (BD Biosciences).

### Direct co-culture

Melanoma cell lines from 5 melanoma patients expressing varying levels of CD9 (as assessed by flow cytometry) were thawed and resuspended in 1 mL of prewarmed PBS + 0.2 µM Cell Trace Violet (1:10,000) for 20 minutes at 37°C. Cells were then washed in RPMI containing 10% heat inactivated fetal bovine serum (FBS) and 1% penicillin-streptomycin. PBMCs from 3 healthy donors were thawed and enriched for NK cells using an EasySep cell isolation kit (17955, EasySep) following the manufacturer’s instructions. Melanoma target cells and enriched NK effector cells were subsequently counted and resuspended at 100,000 cells/100 µL of complete RPMI and plated in 96-well transwell plates with a polycarbonate membrane containing a 0.4 µm pore size (3401, Corning) at a target:effector ratio of 1:1. Target melanoma cells were plated in the lower chamber; effector NK cells were plated in the upper chamber. NK effector cells were plated alone as a negative control. Phorbol 12-myristate 13-acetate (PMA; Sigma-Aldrich) with ionomycin (Sigma-Aldrich) were added to one well at concentrations of 50 ng/mL and 500 ng/mL, respectively. Co-cultures of NK cells with melanoma cell lines were incubated at 37°C for 6 hours. Cells were then collected and transferred to V-bottom 96-well plates for flow cytometry staining and analysis.

### Multiplexed Immunohistochemistry

Sections from formalin-fixed, paraffin-embedded blocks were stained using two Opal multiplexed panels according to the manufacturer’s protocol (PerkinElmer). The first panel was used to screen for NK cells and included the following markers: DAPI, SOX10, CD56, CD16, NKp46, HLA class I, Pan-KIR. The second panel was used to identify dl-NK cells and included the following markers: DAPI, SOX10, CD56, CD16, CD49a, CD9 and VEGF. This staining was performed by the Human Immune Monitoring Shared Resource at CU Anschutz Medical Campus. Slide scanning was performed using the Vectra 3.0 instrument. Multispectral regions of interest were selected and analyzed using inForm Software 3.1.0. Images were spectrally unmixed, evaluated for staining intensities and morphology, tissue segmented based on tissue markers, cell segmented based on nuclear and membrane markers, and phenotypically scored.

### Statistical analysis

Graphs and analyses used GraphPad Prism v11 and RStudio 2026.08.1+195. Distributions were assessed using the D’Agostino–Pearson test. Two-group comparisons used two-tailed unpaired t tests where specified; sex and age comparisons used Wilcoxon rank-sum tests, and melanoma subtype and sampling-site comparisons used Kruskal–Wallis tests. Pearson correlation used log_2_(value + 1) for tumor CD9 expression and dl-NK frequency; Spearman correlation assessed rank association. **Figure 5D** shows a linear fit with a 95% confidence interval. Summary plots show mean ± SEM where indicated. TIL-response analyses and co-culture comparisons without displayed tests were interpreted descriptively. P < 0.05 defined statistical significance.

## ACKNOWLEDGEMENTS

The authors thank the melanoma patients who enrolled in our studies as well as University of Colorado Human Immune Monitoring Shared Resource for performing the multiplex immunohistochemistry staining and the University of Colorado Flow Cytometry Shared Resource for maintaining the Cytek Aurora flow cytometer used in these studies.

## DATA AVAILABILITY

The data generated in this study are available upon request from the corresponding author.

## Synopsis

Decidual-like natural killer (dl-NK) cells promote fetal invasion, immune tolerance, and angiogenesis during early pregnancy, raising the possibility that tumors co-opt this pregnancy-associated program to shape an immune-tolerant microenvironment. Here, we identify dl-NK cells in melanoma for the first time, observe their marked enrichment in a patient with poor response to tumor-infiltrating lymphocyte therapy, and show that direct melanoma–NK cell contact induces CD9 acquisition, a canonical marker of the dl-NK phenotype.

