## Supplementary Files for "Decidual-Like Natural Killer Cells Are Enriched in Melanomas and Acquire CD9 Through Melanoma Cell Contact"

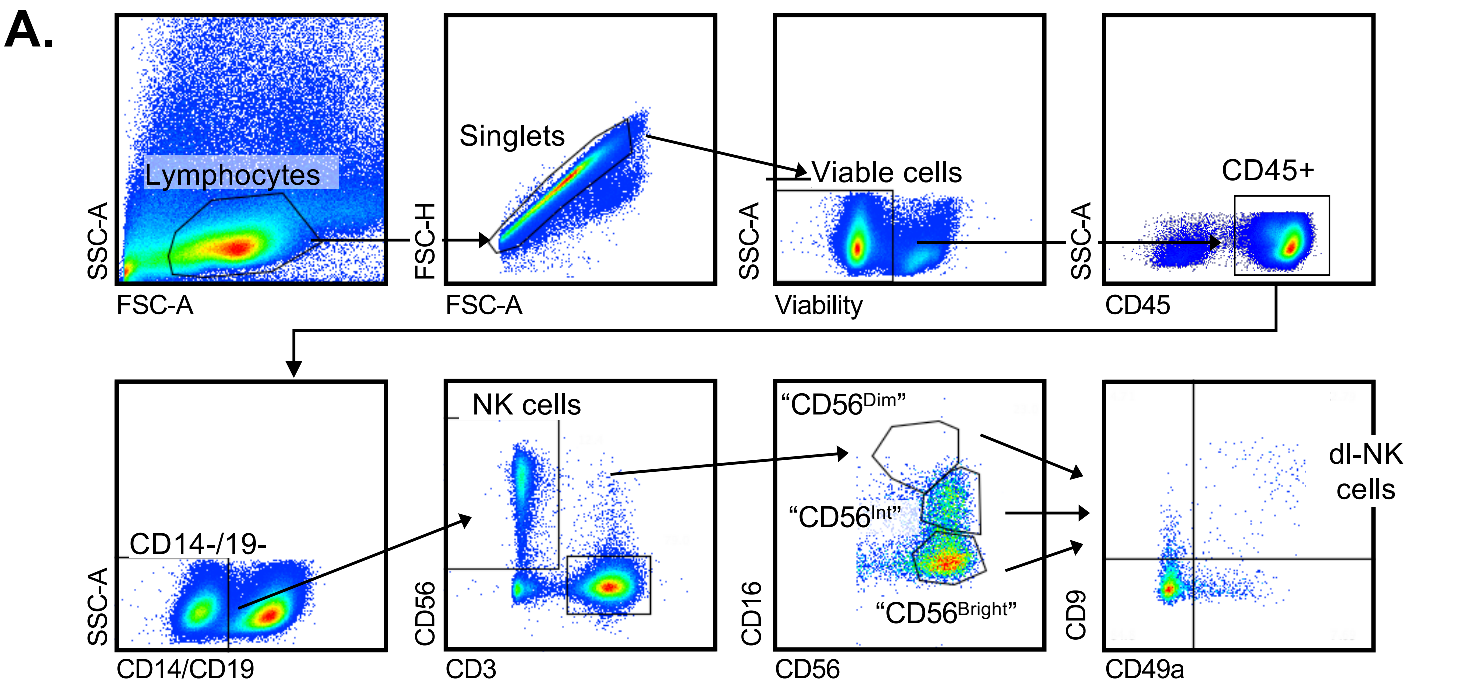


**Supplemental Figure 1. Gating schema used for flow cytometry experiments**

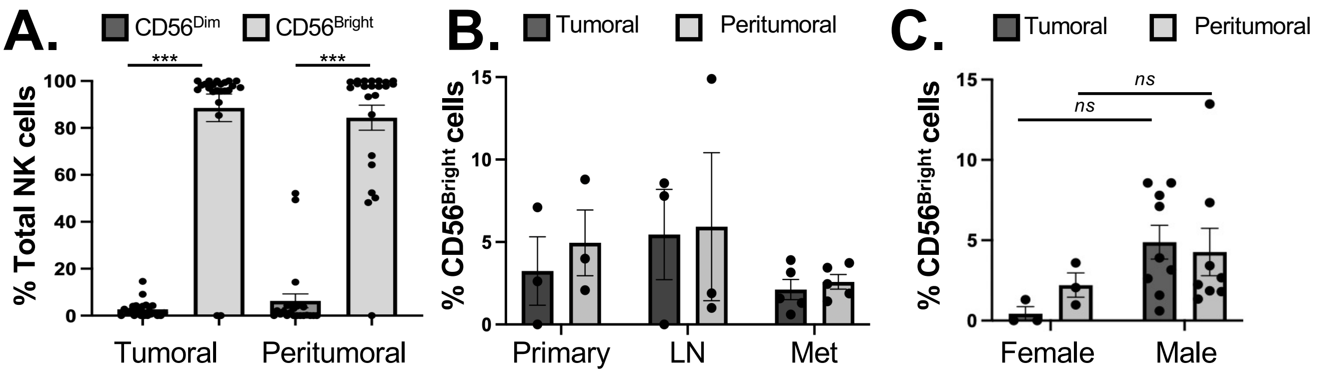


**Supplementary Figure 2: Multiplexed immunohistochemistry show similar distribution in tumoral and peritumoral regions. (A)** Shows quantification $CD56^{Dim}$ ($CD56^{+}CD16^{+}SOX10^{-}$) and $CD56^{Bright}$ ($CD56^{+}CD16^{-}SOX10^{-}$) staining across 23 patient samples within the tumoral and peritumoral regions. (B) shows quantification of dl-NK cells as a percentage of total NK cells within tumoral and peritumoral regions among 3 primary tumors, 3 metastatic lymph nodes and 5 distant metastases. **(C)** Shows quantification of dl-NK cells across the same 11 samples within the tumoral and peritumoral regions divided by gender.

| Supplementary Table 1: Flow Cytometry Antibodies | | | | |
|---|---|---|---|---|
| **Target** | **Fluor** | **Clone** | **Vendor** | **dilution** |
| CD14 | FITC | 61D3 | eBioScience | 1:100 |
| CD14 | RB744 | 61D3 | BD | 1:200 |
| CD19 | FITC | HIB19 | eBioScience | 1:100 |
| CD19 | RB744 | HIB19 | BD | 1:200 |
| CD45 | HI30 | PE-Fire744 | BD | 1:400 |
| CD45 | BV805 | HI30 | BD | 1:300 |
| CD45 | eF506 | HI30 | Invitrogen | 1:100 |
| CD3 | BUV496 | UCHT1 | BD | 1:400 |
| CD8 | BUV395 | RPA-T8 | BD | 1:200 |
| CD56 | BUV737 | NCAM16.2 | BD | 1:400 |
| CD16 | BV605 | 3G8 | BD | 1:1000 |
| CD16 | BV480 | 3G8 | BD | 1:600 |
| CD9 | BV421 | M-L13 | eBioScience | 1:20 |
| CD9 | PE | M-L13 | BD | 1:20 |
| CD9 | SuperBright645 | eBioSN4 | eBioScience | 1:50 |
| CD49a | BB700 | SR84 | BD | 1:100 |
| KIR2DL2/3/S2 | BUV615 | DX27 | BD | 1:200 |
| KIR2DL1/S1 | PE-Cy5.5 | EB6b | Beckman | 1:100 |
| KIR3DL1 | BV786 | DX9 | BD | 1:100 |
| KIR3DL2 | PE | DX31 | Generously provided by UCSF | 1:300 |
| CX3CR1 | BUV805 | 2A9-1 | BD | 1:100 |
| HLA-C | AF488 | DT-9 | R&D | 1:50 |
| NKG2A | APC | Z199 | Beckman Coulter | 1:50 |
| NKG2C | BUV563 | 134591 | BD | 1:50 |
| PD-1 | RB613 | EH12.1 | BD | 1:50 |
| PD-1 | AF647 | EH12.1 | BD | 1:100 |
| TIM-3 | BV711 | 7D3 | BD | 1:300 |
| TIM-3 | PE-CF594 | 7D3 | BD | 1:400 |
| CXCR3 | Pe-Cy7 | 1C6 | BD | 1:100 |

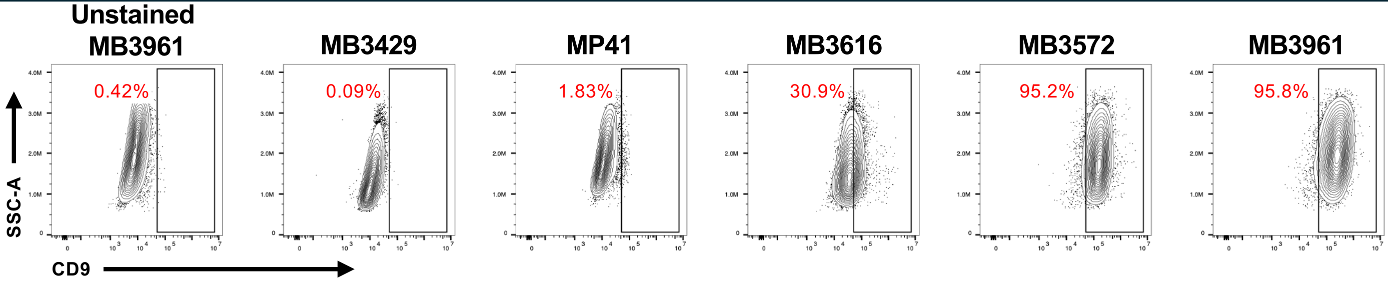


**Supplementary Figure 3: Surface expression of CD9 on melanoma cell lines.** CD9 expression was assessed on 5 unique melanoma cell lines using flow cytometry. MB3429 and MP41 were deemed CD9 negative. MB3616 was deemed CD9+ while MB3572 were labeled as CD9+++ due to very high CD9 surface expression. MB3961 unstained was used to set the CD9 gate.
